# Missing Americans: Early Death in the United States, 1933-2021

**DOI:** 10.1101/2022.06.29.22277065

**Authors:** Jacob Bor, Andrew C. Stokes, Julia Raifman, Atheendar Venkataramani, Mary T. Bassett, David Himmelstein, Steffie Woolhandler

## Abstract

We assessed how many U.S. deaths would have been averted each year, 1933-2021, if U.S. age-specific mortality rates had equaled those of other wealthy nations. The annual number of excess deaths in the U.S. increased steadily beginning in the late 1970s, reaching 626,353 in 2019. Excess deaths surged during the COVID-19 pandemic. In 2021, there were 1,092,293 “Missing Americans” and 25 million years of life lost due to excess mortality relative to peer nations. In 2021, half of all deaths under 65 years and 91% of the increase in under-65 mortality since 2019 would have been avoided if the U.S. had the mortality rates of its peers. Black and Native Americans made up a disproportionate share of Missing Americans, although the majority were White.

**One sentence summary:** In 2021, 1.1 million U.S. deaths – including 1 in 2 deaths under age 65 years – would have been averted if the U.S. had the mortality rates of other wealthy nations.

---

Studies have quantified excess mortality in the United States during the COVID-19 pandemic relative to pre-COVID-19 U.S. death rates.(1–5) However, even before the pandemic, U.S. residents died at younger ages than people in other wealthy nations, particularly from drug overdoses, suicides, and cardiometabolic disorders.(6–12) U.S. life expectancy began diverging from peer countries’ in 1980 and has declined in absolute terms since 2014.(13) In 2018 – prior to COVID-19 – the U.S. suffered 461,000 excess deaths relative to other wealthy countries (17% of all deaths in the U.S. that year),(14) a number that was, coincidentally, similar to the number of U.S. deaths due to COVID-19 each year in 2020 and 2021.(15)

Excess mortality in the U.S. relative to other nations has been linked to structural racism, economic inequality, and underinvestment in public health and social safety net programs in the U.S.(14) These factors, and low vaccine uptake, have also contributed to the severity of the COVID-19 pandemic in the U.S. and its disproportionate toll on racial and ethnic minority individuals and on people with lower income and education levels.(16, 17) These fundamental causes(18) are rendered invisible when excess mortality is computed relative to a U.S. baseline. Additionally, whereas studies of U.S. health disparities typically focus on differences between U.S. racial and ethnic groups, the declining health of White U.S. residents has made it difficult to interpret trends in disparities. External comparisons shed light on the exceptional nature of U.S. mortality trends and the experiences of U.S. racial and ethnic groups.

In this paper, we quantify the number of “Missing Americans” from 1933 through 2021 – the number of deaths that would have been averted each year if U.S. mortality rates had equaled those of other wealthy countries. We extend prior analyses(11, 12, 14, 19) in three directions. First, we contextualize the COVID-19 pandemic within long-term historical trends, computing excess U.S. deaths starting in 1933 (when the U.S. mortality series begins). The longer view paints a clearer picture of the time course of the divergence in mortality between the U.S. and other wealthy nations. It also encompasses World War II, the largest mortality crisis prior to COVID-19 among today’s wealthy nations.(20)

Second, we examine the impact of COVID-19 on the U.S. mortality disadvantage: the number of “Missing Americans” in 2019, on the eve of the COVID-19 pandemic; the change in mortality associated with the COVID-19 pandemic in the U.S. and peer countries; and the impact of the pandemic on the number of “Missing Americans”. We stratify our analyses by age, focusing in particular on adults under 65 years, who experienced rising mortality prior to COVID-19(9, 11), and who are largely excluded from the nearly-universal health coverage and income support available to the elderly, and from social programs targeted to children.

Third, we assess trends in the burden of excess mortality borne by U.S. racial and ethnic groups, given persistent racial disparities in life expectancy,(21) rising mortality among White Americans,(9) and differential mortality impacts of the pandemic.(22, 23)

We conduct each of these analyses using newly-released mortality data through 2021 for the U.S. and other wealthy nations, compiled by the Human Mortality Database (HMD)(24) and the U.S. Centers for Disease Control and Prevention (CDC)(25).

## Methods

We compared mortality trends in the U.S. with mortality trends in 18 other wealthy countries. The comparison set of countries included: Austria, Belgium, Canada, Denmark, Finland, France, Germany, Iceland, Italy, Japan, Luxembourg, Netherlands, Norway, Portugal, Spain, Sweden, Switzerland, and the United Kingdom. These countries represented all those with mortality data available from HMD beginning in 1960 or earlier and ending in 2020 or later, after excluding former-Soviet/Eastern Bloc nations and two countries (Australia, New Zealand) with incomplete data. Our comparator set makes comprehensive use of available data on peer countries, following recent work on life expectancy trends (4, 26). However, we also present “Missing Americans” estimates using two alternate comparators: the other Group of Seven (G7) countries – Canada, France, Germany, Italy, Japan, and the United Kingdom – used in prior work by members of this study team, (14) and the five largest Western European countries – France, Germany, Italy, Spain, UK – used by Preston and Vierboom (11) and Heuveline (19).

Data on deaths and population denominators were obtained from the HMD, which compiles official data from national vital registries and censuses. We augmented the HMD standard long-run mortality series (available from 1960 through 2017 to 2020 for the countries in the sample) with data through 2021 from the HMD’s Short-Term Mortality Fluctuations (STMF) database (both databases were downloaded June 7, 2022). Details on the panel of countries are provided in **Table S1**. All countries were observed starting in 1933 with the exception Portugal (1940), Austria (1947), Japan (1947), Germany (1956), and Luxembourg (1960). The Germany series combines East and West Germany prior to 1989. All countries were observed through 2021, with the exception of Japan, for which we carried forward 2020 ASMRs to 2021.

For each country, we extracted annual age-specific mortality rates, deaths, and denominators (exposure time) for 5-year age groups from the HMD long series. We then extracted data on deaths for the most recent years using the original input data from the STMF. Countries reported deaths for different age bands in the STMF, and we allocated deaths within STMF age groups to 5-year age groups based on the distribution of deaths in the last year available from the HMD long series. Denominators for the most recent years were obtained by fitting a linear trend to the last five years of exposure data available from the HMD long series for each 5-year age group in each country (e.g. Germany, 2013-2017) to obtain predictions for the most recent years. For country-years with overlapping data between the STMF and HMD long series, ASMRs were very similar **(Table S1)**. Finally, to avoid false precision in reporting, we aggregated deaths and exposure time to the ten-year age bands reported by the U.S. in the STMF data: 0-4, 5-14, 15-24, …, 85+ years. For our age-stratified analyses, we constructed six age groups to facilitate presentation: 0-14, 15-44, 45-64, 65-74, 75-84, 85+.

As a benchmark against which to compute “excess” U.S. mortality, we calculated the average ASMR across the other wealthy countries for each age group and year (excluding countries whose data were not available for that year). We weighted countries by population (following (11)), comparing the mortality risks faced by U.S. residents with a representative set of residents of other wealthy nations.

We also compared ASMRs for U.S. racial/ethnic groups relative to the average of other wealthy nations. We obtained data on U.S. mortality and population denominators for 1999-2021, stratified by age and race/ethnicity from the Multiple Cause Mortality Files hosted on the CDC WONDER database (downloaded: April 14, 2022). We computed ASMRs for: Non-Hispanic Black (henceforth Black), Non-Hispanic White (White), Hispanic, Non-Hispanic Asian/Pacific Islander (Asian/Pacific Islander), and Non-Hispanic American Indian/Alaskan Native (Native American) persons. For each population subgroup, ASMRs were extracted for ten-year age groups, aligned with the data from HMD. For ease of exposition, we report on wider age bands: 0-14, 15-44, 45-64, 65-74, 75-84, 85+ years.

### Long-run mortality trends in the U.S. and other wealthy nations

**Figure 1**(**A**) displays long-run trends in age-specific mortality rates for each of the countries in the panel, 1933-2021. Mortality has fallen dramatically in all countries. The early time series bears the imprint of World War II and its aftermath, with high mortality rates observed in Europe and Japan. However, since the 1970s, the position of the U.S. relative to other wealthy nations has progressively deteriorated.

**Fig. 1.**
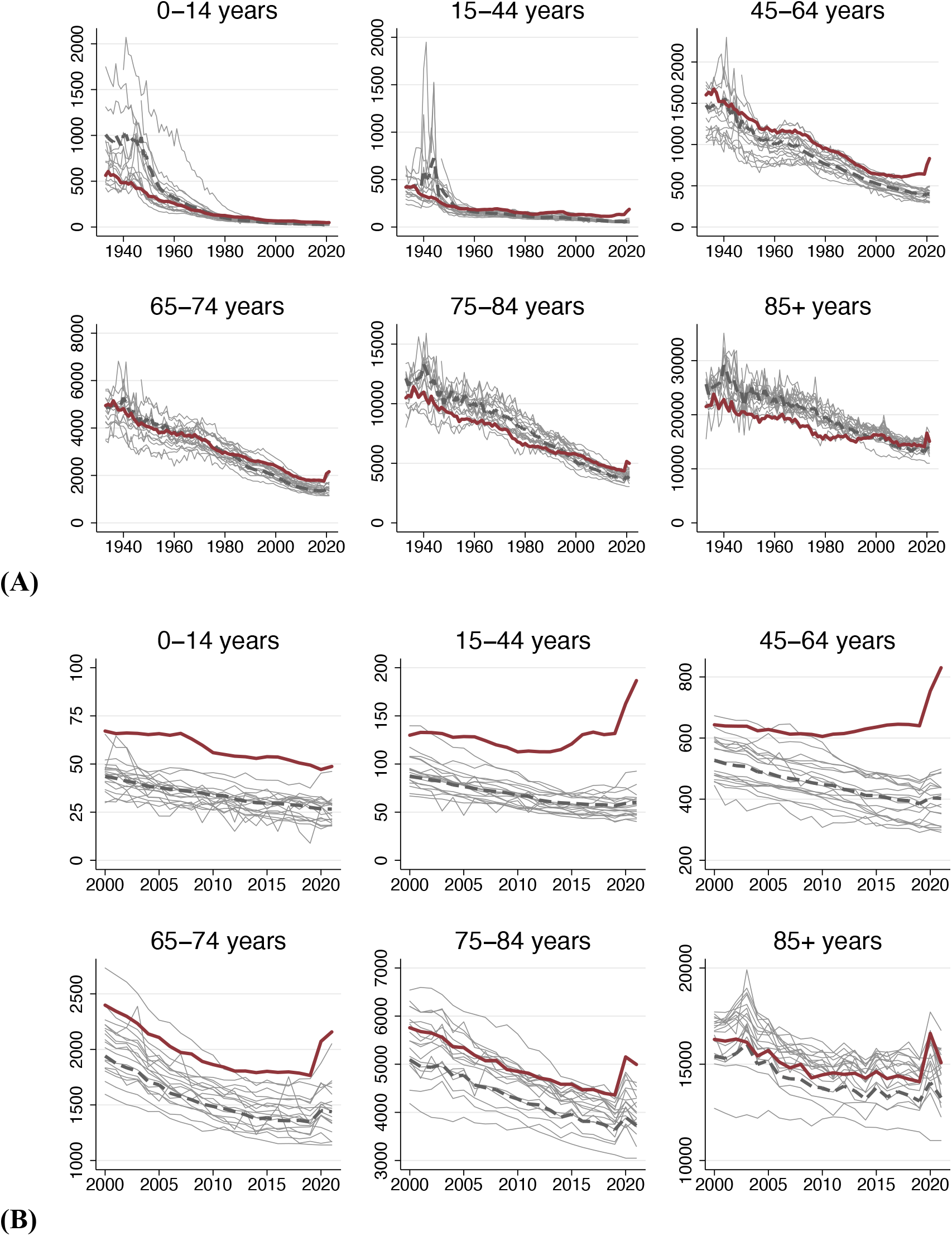
Age-specific mortality trends in the U.S. and other wealthy nations. Source: Human Mortality Database. Note: Figure shows deaths per 100K population, (A) 1933-2021 and (B) 2000-2021. Dark red line is U.S.; pink line is population-weighted average of other wealthy nations; grey lines are country-specific trends for other countries.

**Figure 1**(**B**) zooms in on the same plot for the period 2000-2021. In this period, the gap between the U.S. and other wealthy countries widened, particularly among non-elderly adults (15-64 years). Mortality among U.S. non-elderly adults increased in absolute terms throughout this period. The COVID-19 pandemic appears as a spike in mortality in 2020 and 2021. However, the size of the 2020 increase differed by country and age group. From 2020 to 2021, U.S. mortality rates declined in older age groups (over 75 years - a group that had experienced a large increase between 2019 and 2020) but increased among younger age groups. **Figure S1** summarizes ASMR trends across age groups, standardizing by the 2000 U.S. population.

**Figure S2** displays trends in the ratio of U.S. mortality to the average of other wealthy countries for each age group, 1933-2021. The U.S. mortality advantage in the late 1930s and 1940s reflects the disproportionate mortality shock of WW II on younger age groups in many of the comparison countries. In general, however, younger U.S. age groups had elevated mortality rates relative to peer nations, while older U.S. age groups had lower mortality. This pattern began to shift in the late 1970s as the U.S. advantage at older ages disappeared. Since 2000, trends in rate ratios for different age groups have diverged, increasing rapidly for persons under 65 years, while remaining approximately stable (until the onset of the COVID-19 pandemic) for the older age group.

How many U.S. deaths would have been averted each year if the U.S. had ASMRs equal to the average of other wealthy nations? As a descriptive counterfactual, we computed the number of deaths that would have occurred in the U.S. if the U.S. population had experienced the age-specific mortality profile of peer nations. We then subtracted this number from the actual number of U.S. deaths at each age to compute the number of “Missing Americans”.

**Figure 2A** shows, for each year 1933-2021, the number of deaths that occurred in the U.S. and the number that would have occurred if the U.S. had the ASMRs of other wealthy nations. **Figure 2B** shows the number of “Missing Americans” each year. Since the mid-1970s, the U.S. has experienced a steady and approximately linear increase in the number of excess deaths relative to other wealthy nations. The rising number of Missing Americans was driven by increases in excess mortality over 65 years from the late 1970s through the 1990s and increases in excess mortality under 65 years since 2000 (**Figure S3**). We discuss changes during the COVID-19 pandemic below.

**Fig. 2.**
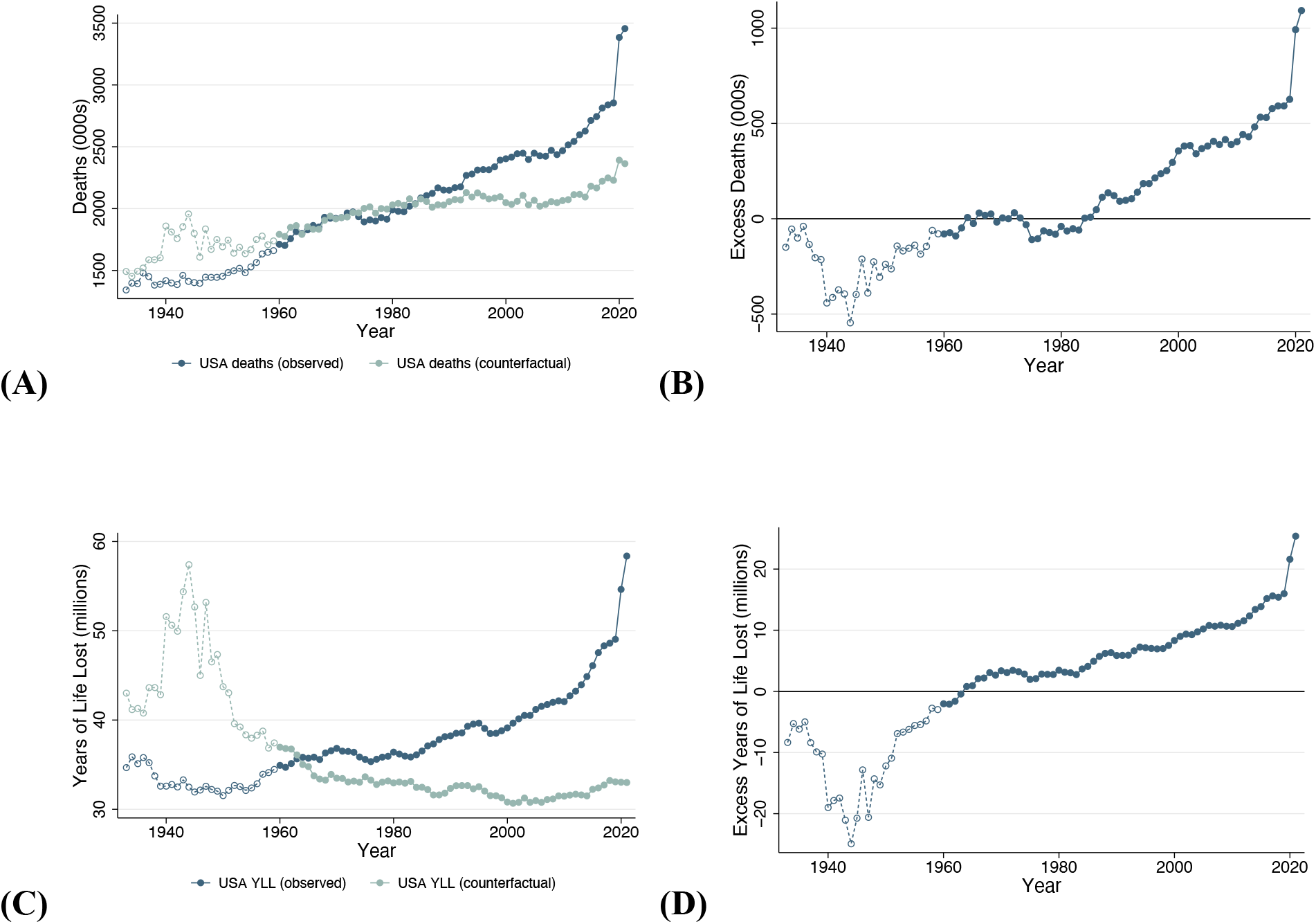
Excess deaths and years of life lost in the U.S. relative to other wealthy nations, 1933-2021. Source: Human Mortality Database. Note: Figure shows number of deaths in the U.S. and the number of deaths that would have occurred in the U.S. if the U.S. had age-specific mortality rates equal to the average of other wealthy nations. The average of other wealthy nations excludes Portugal prior to 1940, Austria and Japan prior to 1947, Germany prior to 1956, and Luxembourg prior to 1960. From 1960, all countries are represented. Panel (A) shows deaths in the U.S. and counterfactual; panel (B) shows the difference between the two; panels (C) and (D) show analogous plots for years of life lost (YLL), where each death is weighted by U.S. age-specific life expectancy in the year it occurred.

The increase in excess U.S. mortality is even more stark when using a “years of life lost” (YLL) metric. Rather than treating every death the same, YLL weights deaths by the number of years that a person would have been expected to live had they survived. Following (11), we compute YLL by weighting each death by the U.S. age-specific life expectancy in that year. **Figure 2C** shows YLL associated with observed U.S. deaths and YLL if the U.S. had ASMRs of its peers. **Figure 2D** shows the difference in YLL associated with excess U.S. deaths. Whereas U.S. YLL have increased continuously since 1960, YLL would have flatlined since 1960 if the U.S. had the ASMRs of other wealthy nations. The rise in excess deaths and YLL were not explained by population aging, as shown in **Figure S4**, which presents age-standardized trends.

In 2019, on the eve of the COVID-19 pandemic, the U.S. experienced 2,854,820 deaths. If the U.S. had ASMRs of the other wealthy countries, there would have been 2,228,467 deaths, a difference that represents 626,353 excess deaths (**Table 1, Figure 2**). Over half (51%) of these excess U.S. deaths were in persons younger than 65, a group that incurred just 26% of all U.S. deaths. Forty-three percent of U.S. deaths in 2019 under 65 years and 15% of deaths over 65 years would have been avoided if U.S. mortality rates had equaled the average of other wealthy nations (**Table 1, Table S2**).

**Table 1.**
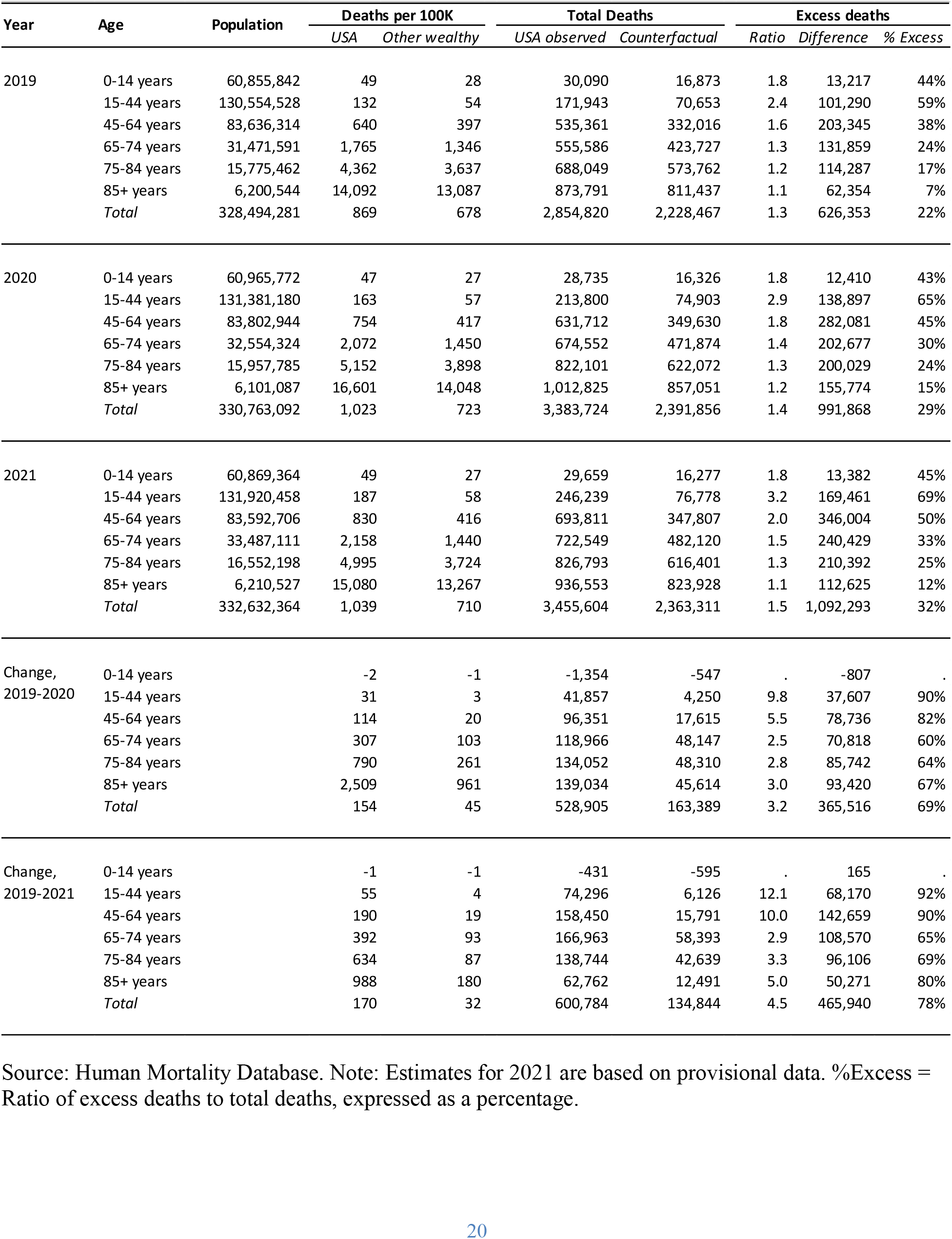
Excess mortality in the U.S. relative to other wealthy nations: 2019-2021.

**Figure 3** presents detailed age-profiles of U.S. excess mortality for selected years: 1933, 1960, 1980, 2000, 2019-2021. The figure displays mortality rate ratios, excess deaths, and years of life lost in the U.S. relative to other wealthy nations. The divergence of the U.S. from peer countries occurred at different points in time for different age groups. From 1933 to 1960 (bookending WWII and its aftermath), U.S. mortality rates were similar to those in other nations. From 1960 to 1980, mortality in the U.S. relative to peers increased for younger age groups (15-34 years). From 1980 to 2000, U.S. mortality increased relative to peers for older age groups (65+ years). From 2000 to 2019, U.S. mortality increased relative to peers among younger and middle-aged adults. **Figure 3** highlights the extent to which the divergence in mortality has accelerated in the 21^st^ century. For many age groups, the difference in excess deaths between 2000 and 2019 exceeded the change from 1933 to 2000. In 2019, mortality rate ratios comparing the U.S. to the average of other wealthy nations were highest among younger adults; older adults experienced the greatest number of excess deaths; and middle-aged adults the largest number of YLL.

**Fig. 3.**
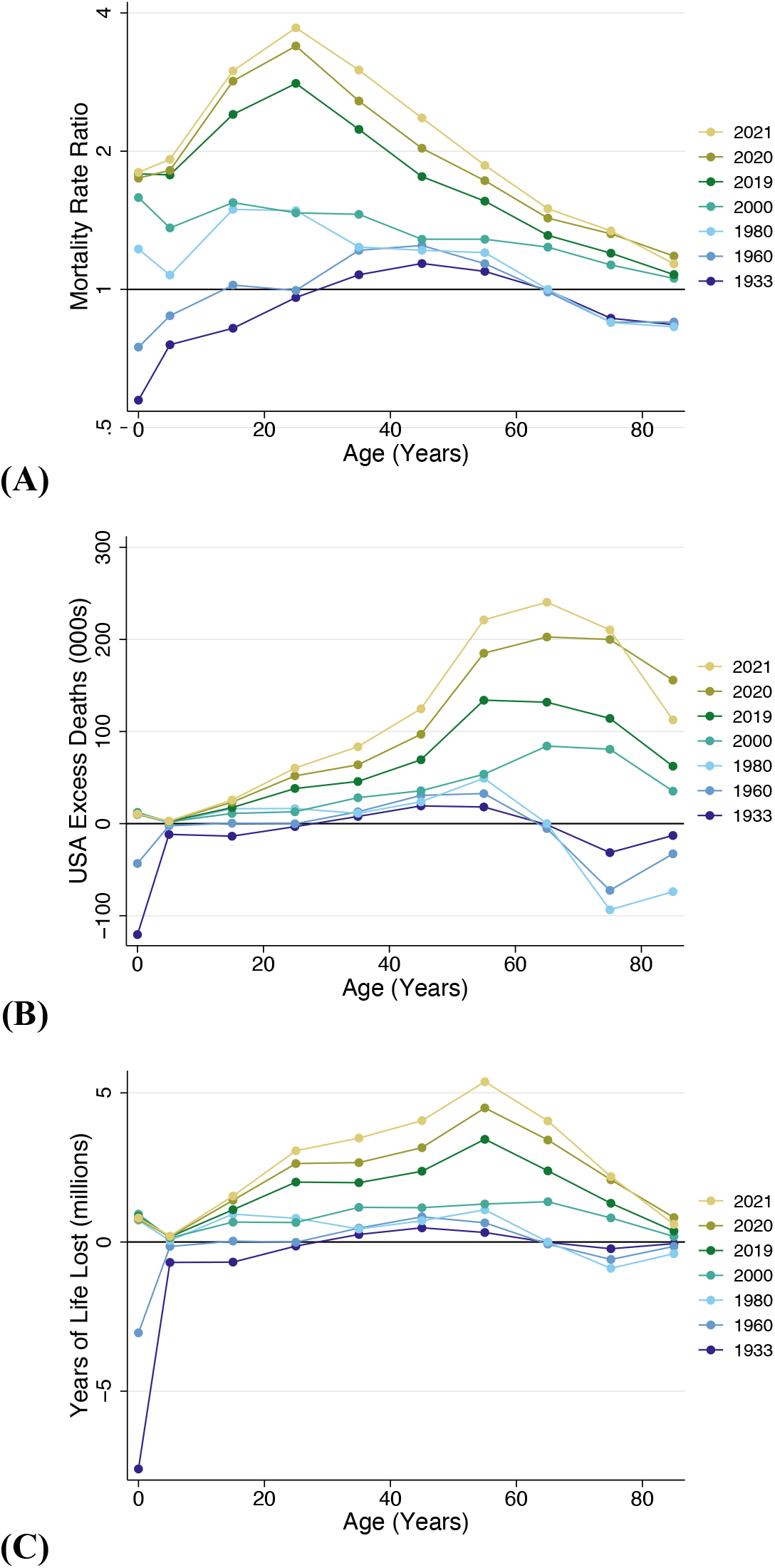
Excess mortality by age group for select years: 1933 – 2021. Source: Human Mortality Database. Note: Figure shows (A) mortality rate ratios, (B) excess deaths, and (C) years of life lost in the U.S. relative to what would have occurred if the U.S. had age-specific mortality rates equal to the average of other wealthy nations. The group of other wealthy nations excludes Portugal, Austria, Japan, and Germany in 1933. From 1960, all countries are represented. Age groups are: 0-4, 5-14, 15-24, …, and 85+ years.

### Impact of COVID-19 pandemic on the U.S. mortality disadvantage

The last two years of our mortality series reflect the toll of the COVID-19 pandemic, which in 2020 caused the largest single-year increase in mortality for the U.S. and many other wealthy countries since World War II. From 2019 to 2020 and 2021, mortality increased in all US age groups except children 0-14 years (**Figure 1, Table 1**).

Across countries, however, the toll of the pandemic was uneven. To assess the impact of COVID-19, we compared changes in mortality from 2019 to 2020 and 2021 in the U.S. relative to other wealthy nations. In the U.S., the pandemic was associated with an increase of 528,905 deaths between 2019 and 2020 and an increase of 600,784 deaths between 2019 and 2021. If the U.S. had ASMRs equal to the average of other wealthy nations, then 69% of the 2020 increase and 78% of the 2021 increase would have been avoided (**Table 1, Table S1**).

During the COVID-19 pandemic, the number of Missing Americans grew from 626,353 in 2019 to 991,868 in 2020, and to 1,092,293 in 2021 (**Table 1, Table S1, Figure 2**). In 2021, 48% of these excess deaths occurred before age 65. Annual YLL associated with the U.S. mortality disadvantage increased from 16.0 million in 2019 to 25.4 million in 2021, up from 8.3 million in 2000 (**Figure 2**).

The COVID-19 pandemic took a particularly large toll among younger people in the U.S., far greater than in other wealthy nations (**Figure 1**). Among people under 65 years, the 2019-2020 increase in mortality was five times greater in the U.S. than in other wealthy nations, and the 2019-2021 increase in mortality was ten times greater in the U.S. If the U.S. had the ASMRs of its peers, 84% of the 2019 to 2020 increase in U.S. mortality under age 65 and 91% of the 2019 to 2021 increase in U.S. mortality under age 65 would have been avoided (**Table 1, Table S1**). Interestingly, other wealthy nations saw under-65 mortality hold steady at 160 deaths per 100K from 2020 to 2021; in the U.S., under-65 mortality increased, from 317 to 351 deaths per 100K. Estimates of U.S. excess deaths in 2019-2021 were qualitatively similar when using alternative international comparison sets (**Table S3**).

### Who are the missing Americans? Excess mortality by U.S. race/ethnicity

**Figure 4A** presents ratios comparing ASMRs for each U.S. race/ethnicity to the average of other wealthy countries. Data are presented for four years -- 1999, 2019, 2020, and 2021 – enabling assessment of trends in excess mortality by race/ethnicity both before and during the COVID-19 pandemic. The figure shows important commonalities across U.S. racial/ethnic groups. Mortality rate ratios relative to other nations increased over time for all race/ethnicities. Mortality rate ratios were largest – for all groups – among young adults (15-44 years).

**Fig. 4.**
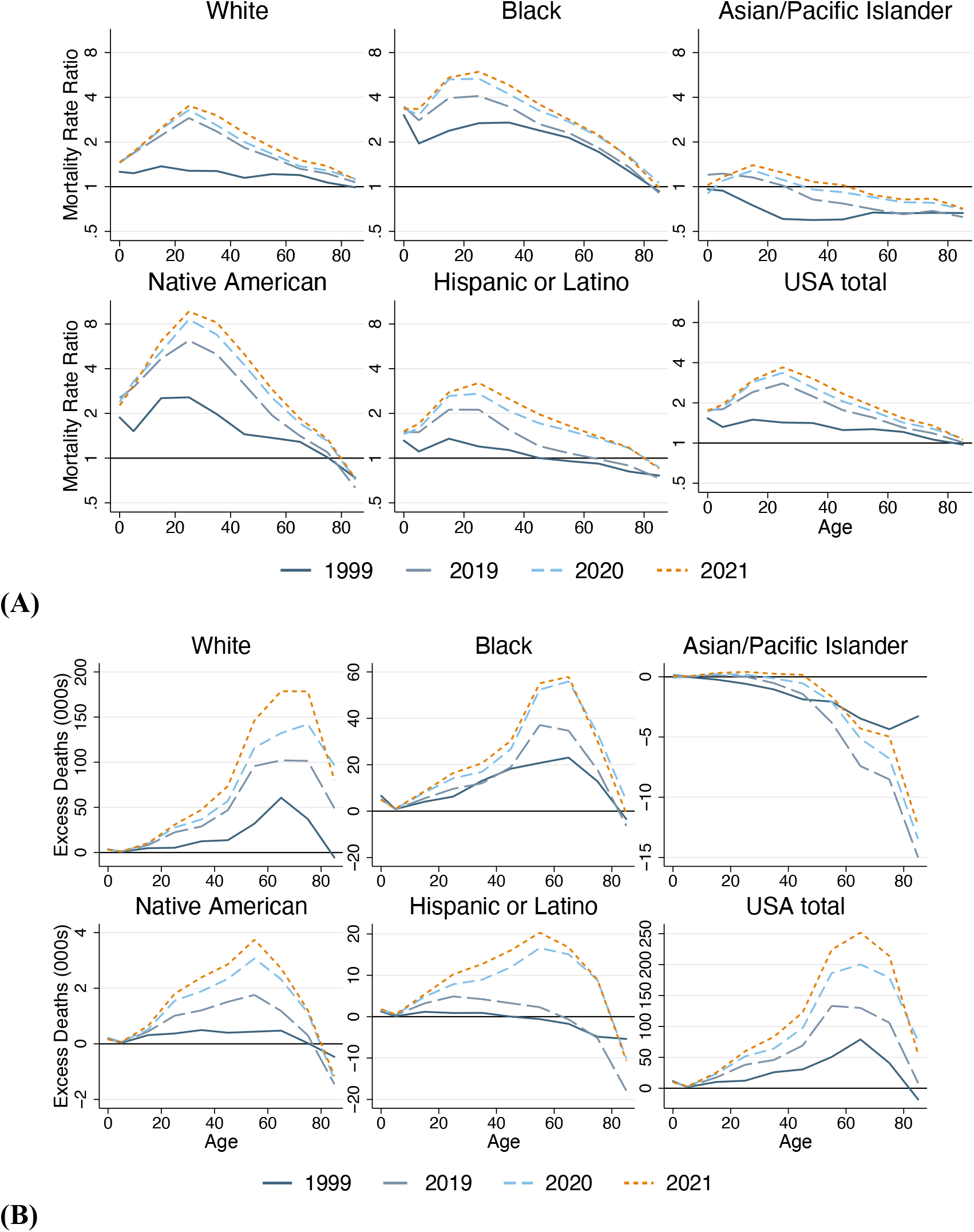
Excess mortality by U.S. race/ethnicity relative to the average of other wealthy nations, 1999-2021. Source: Multiple Cause Mortality Files from CDC Wonder and Human Mortality Database. Figure shows (A) mortality rate ratios and (B) number of excess deaths for each group, stratified by age: 0-14, 15-44, 45-64, 65-74, 75-84, 85+ years. U.S. racial/ethnic groups are: Hispanic, Non-Hispanic Black (Black), Non-Hispanic White (White), Non-Hispanic Asian/ Pacific Islander (Asian/Pacific Islander), Non-Hispanic American Indian/Alaskan Native (Native American).

However, there were also key differences in levels and trends. In 2019, Black and Native American U.S. residents aged 15-44 years had mortality rates 5.3 times and 3.8 times higher respectively than the average of other wealthy countries. Mortality among White and Hispanic U.S. residents ages 15-44 years was also elevated at 2.5 times and 1.8 times the average of other nations, respectively. By contrast, in 2019 Asian/Pacific Islander U.S. residents had mortality rates below the average of other nations. White and Native Americans experienced the largest increases in mortality from 2000 to 2019.

During the COVID-19 pandemic, mortality rose for all U.S. racial/ethnic groups in all adult age groups (**Figure 3A, Figure 5**). Native American, Black, and Hispanic U.S. residents experienced the largest increases, and non-elderly adults of these racial and ethnic groups experienced the largest relative increases compared to other wealthy nations. For example, from 2019 to 2021, age-standardized, under-65 mortality increased by 249 deaths per 100K among Native American residents, 140 deaths per 100K among Black residents, 108 deaths per 100K among Hispanic residents, 71 deaths per 100K among White residents, and 35 deaths per 100K among Asian residents – compared to an increase of just 7 deaths per 100K in other wealthy nations (**Figure S5**). The increase in Hispanic Americans’ death rates largely erased their previous survival advantage relative to non-Hispanic White Americans and pushed their mortality rates above the average of other wealthy countries at all but the oldest ages.

**Fig. 5.**
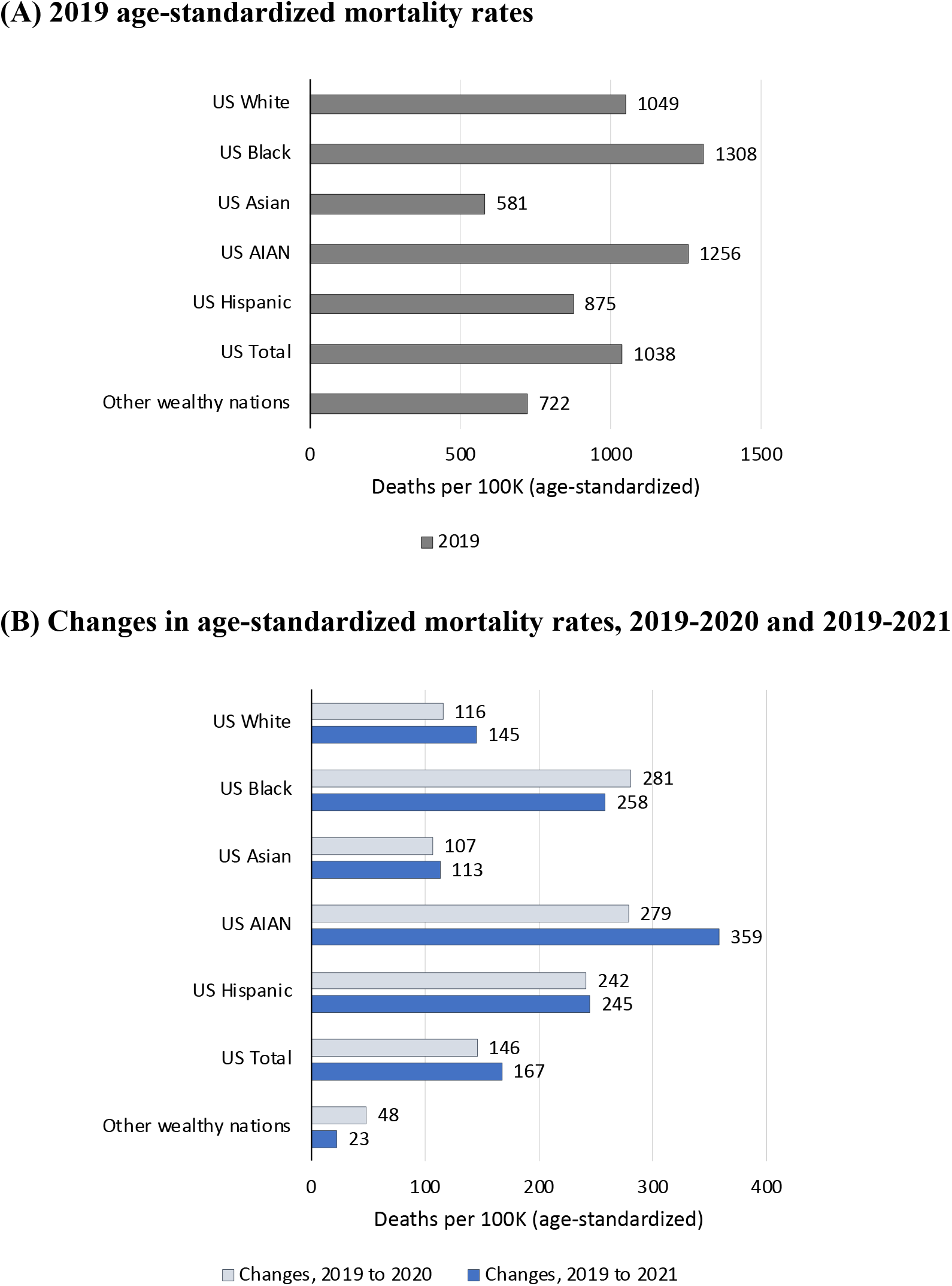
Age-standardized mortality rates for U.S. racial and ethnic groups and the average of other wealthy nations: 2019-2021. Source: Multiple Cause Mortality Files from CDC Wonder and Human Mortality Database. Note: Figure shows (A) 2019 mortality rates and (B) changes in mortality 2019 to 2020 and 2019 to 2021. Mortality rates are standardized to the 2019 U.S. population age distribution. Figure compares mortality rates for U.S. racial/ethnic groups: Hispanic, Non-Hispanic White (White), Non-Hispanic Black (Black), Non-Hispanic Asian/Pacific Islander (Asian), Non-Hispanic American Indian/Alaskan Native (AIAN).

**Table 2** and **Figure 3B** show the number of “Missing Americans” by race/ethnicity, comparing each U.S. racial and ethnic group to the average of other wealthy nations. Despite relatively higher mortality rates among Black and Native Americans, and larger mortality impacts of COVID-19 on Black, Native and Hispanic Americans, most “Missing Americans” were White, a consequence of the larger and much older population of White U.S. residents. In 2021, there were 749,752 excess deaths among White Americans (who represent 61% of the population) and 224,197 excess deaths among Black Americans (13% of the population). There were 82,051 excess deaths among Hispanic Americans, who have a much younger age distribution than other groups despite representing 19% of the U.S. population. There were 14,423 excess deaths among Native Americans, who represent 1% of the U.S. population. Finally, U.S. residents identifying as Asian or Pacific Islander had lower mortality than the average of other wealthy nations and - 22,085 excess deaths (**Table 2**).

**Table 2.** Excess deaths by U.S. race/ethnicity relative to other wealthy nations: 2019-2021.

| Race/<br>ethnicity | Age | Population<br>(2019) | 2019 |  |  | 2020 |  |  | 2021 |  |  |
| --- | --- | --- | --- | --- | --- | --- | --- | --- | --- | --- | --- |
|  |  |  | Deaths | Excess | % Excess | Deaths | Excess | % Excess | Deaths | Excess | % Excess |
| White | 0-14 | 31,524,362 | 13,076 | 4,316 | 33% | 12,528 | 4,196 | 33% | 12,566 | 4,231 | 34% |
|  | 15-44 | 72,844,178 | 99,126 | 59,375 | 60% | 115,919 | 74,054 | 64% | 131,061 | 88,424 | 67% |
|  | 45-64 | 55,142,376 | 367,261 | 142,602 | 39% | 405,621 | 173,189 | 43% | 450,505 | 219,944 | 49% |
|  | 65-74 | 23,716,452 | 421,407 | 102,093 | 24% | 485,693 | 132,130 | 27% | 529,785 | 178,608 | 34% |
|  | 75-84 | 12,436,703 | 553,760 | 101,430 | 18% | 639,678 | 142,157 | 22% | 653,677 | 178,396 | 27% |
|  | 85+ | 5,241,403 | 734,885 | 48,967 | 7% | 831,544 | 96,280 | 12% | 774,513 | 80,121 | 10% |
|  | Total | 200,905,474 | 2,189,515 | 458,784 | 21% | 2,490,983 | 622,007 | 25% | 2,552,108 | 749,725 | 29% |
| Black | 0-14 | 9,227,154 | 8,615 | 6,038 | 70% | 8,387 | 5,851 | 70% | 8,490 | 5,953 | 70% |
|  | 15-44 | 18,703,220 | 36,962 | 27,110 | 73% | 49,524 | 39,086 | 79% | 56,259 | 45,628 | 81% |
|  | 45-64 | 10,248,785 | 96,151 | 56,080 | 58% | 121,202 | 79,103 | 65% | 127,167 | 85,418 | 67% |
|  | 65-74 | 3,149,591 | 77,055 | 34,650 | 45% | 103,700 | 55,890 | 54% | 105,377 | 57,890 | 55% |
|  | 75-84 | 1,387,735 | 68,321 | 17,848 | 26% | 89,555 | 33,595 | 38% | 83,760 | 30,301 | 36% |
|  | 85+ | 519,318 | 61,638 | -6,323 | -10% | 79,412 | 4,571 | 6% | 69,688 | -992 | -1% |
|  | Total | 43,235,803 | 348,742 | 135,404 | 39% | 451,780 | 218,096 | 48% | 450,741 | 224,197 | 50% |
| Asian/PI | 0-14 | 3,553,388 | 1,180 | 202 | 17% | 995 | -68 | -7% | 1,116 | 52 | 5% |
|  | 15-44 | 9,495,354 | 4,940 | -378 | -8% | 5,911 | 281 | 5% | 6,716 | 983 | 15% |
|  | 45-64 | 5,076,497 | 13,881 | -5,213 | -38% | 17,700 | -2,613 | -15% | 18,668 | -1,470 | -8% |
|  | 65-74 | 1,582,888 | 13,899 | -7,413 | -53% | 18,889 | -5,134 | -27% | 19,554 | -4,306 | -22% |
|  | 75-84 | 747,503 | 18,673 | -8,514 | -46% | 23,982 | -6,792 | -28% | 24,438 | -4,960 | -20% |
|  | 85+ | 304,936 | 24,899 | -15,007 | -60% | 32,096 | -13,477 | -42% | 30,656 | -12,384 | -40% |
|  | Total | 20,760,566 | 77,472 | -36,323 | -47% | 99,573 | -27,802 | -28% | 101,148 | -22,085 | -22% |
| AIAN | 0-14 | 604,145 | 439 | 272 | 62% | 392 | 239 | 61% | 371 | 217 | 59% |
|  | 15-44 | 1,177,885 | 3,271 | 2,655 | 81% | 4,600 | 3,948 | 86% | 5,494 | 4,830 | 88% |
|  | 45-64 | 650,645 | 5,855 | 3,272 | 56% | 8,117 | 5,412 | 67% | 9,289 | 6,606 | 71% |
|  | 65-74 | 212,383 | 4,041 | 1,182 | 29% | 5,551 | 2,318 | 42% | 5,927 | 2,716 | 46% |
|  | 75-84 | 90,180 | 3,589 | 309 | 9% | 4,842 | 1,123 | 23% | 4,769 | 1,216 | 25% |
|  | 85+ | 30,205 | 2,501 | -1,452 | -58% | 3,253 | -1,290 | -40% | 3,129 | -1,162 | -37% |
|  | Total | 2,765,443 | 19,696 | 6,238 | 32% | 26,755 | 11,750 | 44% | 28,978 | 14,423 | 50% |
| Hispanic | 0-14 | 15,661,797 | 6,560 | 2,180 | 33% | 6,220 | 2,003 | 32% | 6,555 | 2,337 | 36% |
|  | 15-44 | 28,066,338 | 27,252 | 12,327 | 45% | 37,364 | 21,547 | 58% | 44,422 | 28,309 | 64% |
|  | 45-64 | 12,205,136 | 50,104 | 5,555 | 11% | 76,551 | 28,551 | 37% | 83,933 | 36,353 | 43% |
|  | 65-74 | 2,822,119 | 37,337 | -659 | -2% | 58,239 | 15,064 | 26% | 59,614 | 16,731 | 28% |
|  | 75-84 | 1,307,751 | 42,375 | -5,189 | -12% | 62,231 | 8,885 | 14% | 59,895 | 8,934 | 15% |
|  | 85+ | 509,096 | 48,759 | -17,864 | -37% | 65,092 | -10,030 | -15% | 60,334 | -10,612 | -18% |
|  | Total | 60,572,237 | 212,387 | -3,650 | -2% | 305,697 | 66,020 | 22% | 314,753 | 82,051 | 26% |

Source: Public-use Multiple Cause Mortality Files from CDC Wonder and Human Mortality Database. Note: Estimates for 2021 are based on provisional data. Table displays rates for U.S. racial/ethnic groups: Hispanic, Non-Hispanic White (White), Non-Hispanic Black (Black), Non-Hispanic Asian/Pacific Islander (Asian), Non-Hispanic American Indian/Alaskan Native (AIAN).

## Discussion

The number of “Missing Americans” increased sharply during the COVID-19 pandemic, exacerbating large pre-existing disparities in early death between the U.S. and other wealthy nations. In 2019, 626,000 U.S. deaths would have been averted had U.S. mortality rates equaled the average of peer nations. Excess U.S. deaths increased to 1,092,000 in 2021. In 2020 and 2021, more than half of all deaths under 65 years would have been averted if the U.S. had experienced the mortality rates of its peers. The 1.1 million “Missing Americans” in 2021 would have enjoyed an estimated 25 million additional years of life had they survived.

Our findings are consistent with recent reports indicating a growing life expectancy gap between the U.S. and other countries.(4, 20, 26–28) Using a similar set of comparator countries, Woolf and colleagues estimated that the U.S. life expectancy deficit increased from 1.9 years to 3.1 years between 2010 and 2018, and widened further to 4.7 years in 2020(4) and 5.3 years in 2021 (26). While period life expectancy is a widely used summary measure of mortality, it may be misinterpreted as reflecting mortality differences at end of life rather than at younger ages where the greatest differences lie between the U.S. and peer nations.

“Missing Americans” offers an alternative, easily interpretable metric: the number of U.S. deaths that would have been averted if the U.S. had the age-specific mortality rates of its peers. Our analysis builds on our (14) and others’ (11, 12) previous analyses documenting excess U.S. deaths prior to COVID-19 and during the pandemic (19), as well as work on racial disparities in excess deaths before (14, 29) and during the COVID-19 pandemic (22, 23, 30).

Other studies have computed “excess deaths” relative to different benchmarks. Our choice of an international comparator offers insights not available from comparison groups within the U.S. For example, studies computing excess mortality relative to pre-COVID-19 trends within the same country take pre-pandemic mortality as given.(1) However, U.S. mortality has diverged from other wealthy nations since the 1970s, after tracking close to other nations for most of the post-World War II period. As we show, there were half a million Missing Americans in 2019 even before COVID-19. Other studies have compared mortality rates across racial/ethnic or socioeconomic groups within the U.S.(22) However, life expectancy of White Americans – particularly those with less than a college degree and those residing in non-metro areas – has stagnated or fallen since the 1990s. Using White Americans as a benchmark renders invisible the mortality crisis among White U.S. residents and underestimates excess mortality for other U.S. racial and ethnic groups.

About half of the Missing Americans died before reaching age 65. Our analysis quantifies the large and widening gap in mortality under 65 years between the U.S. and other wealthy nations, a gap that predated COVID-19 but substantially increased during the pandemic. In 2019, U.S. residents under age 65 years had mortality rates 76% higher than non-elderly residents of other wealthy nations. This number increased to 98% in 2020 and 120% in 2021. Ninety-one percent of the increase in under-65 mortality in the U.S. from 2019 to 2021 would have been averted if the U.S. had the age-specific mortality rates of its peers. Deaths among younger adults imply larger numbers of YLL than deaths among older adults. Additionally, deaths to working-age adults have social consequences, including psychological trauma and economic precarity for surviving children, other relatives, and community members.(31–34)

Although most Missing Americans were White, Black and Native Americans had higher mortality rates than White Americans prior to the pandemic, and Black, Native, and Hispanic Americans had larger mortality increases during the pandemic. Nevertheless, White Americans had mortality rates substantially higher than the average of other wealthy nations, particularly for working age adults. And with their much older age distribution and larger population than other U.S. racial and ethnic groups, White Americans accounted for 69% of excess deaths in 2021. Black Americans accounted for 21%. The underperformance of the U.S. with respect to peer nations is a multiracial phenomenon.

In 2020 and 2021, COVID-19 joined firearm injuries, car accidents, drug overdoses, and cardiometabolic diseases (12) as proximate reasons why U.S. residents die younger than residents of other wealthy nations. However, the exceptionally large spike in early death in the U.S. during 2020-2021 was not only due to the emergence of a novel pathogen. Other nations were similarly exposed. Rather, the mortality impact of the pandemic in the U.S. was a product of longstanding social inequities and policy failures that made the U.S. particularly vulnerable, factors that had contributed to the U.S. mortality disadvantage before the pandemic. Beginning with the genocide of native populations and the enslavement of people of African descent,(35, 36) and continuing through a long history of discrimination, segregation, and exclusion, U.S. policies have directly harmed the health of Black and Native Americans (21), and these harms have reverberated through the intergenerational transfer of resources, power, and health.

In recent decades, policy failures have adversely affected the health of U.S. residents of all racial/ethnic groups, particularly those without a college degree.(37) Since the 1970s, the U.S. – like other wealthy countries – has undergone structural economic changes, with increased exposure to trade,(38) automation,(39) and a shift to service sector employment. The U.S. has failed to protect less-educated workers from the adverse health consequences of these changes. Stagnant minimum wages and losses of collective bargaining protections(40) have contributed to widening economic inequality. A scant safety net for working age adults and the absence of universal healthcare have privatized risk, tying health more closely to personal wealth and employment.(41) Additionally, lax regulation of opioids, firearms, environmental pollutants, unhealthy foods, and workplace safety have contributed to elevated U.S. mortality, particularly among lower-educated and lower-income people. Increasingly divergent policies at the state-level have resulted in widening health gaps.(42) Ironically, in those geographic areas of the U.S. where excess mortality has increased the most, voters have turned towards policy-makers who have further undermined population health(43, 44), e.g. through refusal to expand Medicaid or to implement firearm regulations.(45) Underlying these policy trends, the commitment of a majority of U.S. White residents to a political program of deregulation and divestment from the social determinants of health has hamstrung government efforts to protect the health of all U.S. residents, inflicting the greatest harm on the most vulnerable.(46)

During COVID-19, these same “fundamental causes” – socioeconomic inequality, a limited safety net, and a politics unaccountable to population health – manifested in a large increase in excess U.S. deaths. Residential segregation, limited ability to work from home, and distrust of the medical establishment contributed to excess risk among low-income and minority populations (30, 47). Gaps in the safety net forced many low-income people to work despite high infection risk.(17) Housing insecurity increased crowding and transmission at home. The exclusion of immigrant populations from the safety net(48), anemic workplace protections(49), and crowded housing(50) contributed to the large impact of COVID-19 among U.S. Hispanic residents.(51) And failure to mandate that employers give their workers time off to get vaccinated contributed to disparities in uptake.(52) Finally, the politicization of the pandemic – including mask use, physical distancing, and vaccine uptake – by right-wing policy-makers and media have resulted in substantial excess COVID-19-related mortality and contagion, particularly among U.S. White residents and those with less than a college education.(53)

How can the U.S. reduce the number of “Missing Americans” lost to early death each year? Expanding current U.S. policies with demonstrated health impact – and ensuring that all U.S. residents have access to health care and public benefits – is a place to start. Recent evidence indicates that Medicaid improves economic security and reduces mortality;(54, 55) that minimum wage increases and EITC expansion reduce suicide;(56) that unemployment insurance subsidies improve food security;(57) that childhood programs (e.g. access to nutritional assistance(58) and early childhood education(59)) have health impacts and other benefits many times the initial investment; and that firearm(60) and environmental regulation(61) works. Learning from peer nations, the U.S. could extend social protections and universal health coverage to non-elderly adults and enhance the regulation of pollutants and firearms.

Our analysis has limitations. First, the mortality data for 2021 are preliminary due to the lag time between the occurrence and registration of deaths. Still, in previous years, by 26 weeks after the end of the year (similar to the time frame when we downloaded the data), 99% of U.S. deaths have been recorded(62). Data for Japan were missing for 2021 and were imputed using 2020 death rates. Second, differential reporting of U.S. race and ethnicity on death certificates and in Census denominators may bias estimates. In particular, American Indian/Alaskan Native “race” and Hispanic “ethnicity” are underreported on death certificates, which may lead to underestimation of deaths for these groups (63, 64). Third, the literature on U.S. mortality in international context has used a range of country comparison sets and results vary depending on the countries included. We utilized data on all other wealthy countries with a lengthy time series, excluding former communist countries. However, our findings are robust to alternate comparison sets used in other studies of excess deaths (11, 14).

It is unknown whether U.S. excess deaths will persist at 2021 levels into the future, or if the number of Missing Americans will revert towards pre-COVID levels. Underlying factors for high COVID-19 mortality, such as low vaccination rates; high rates of diabetes, obesity, and hypertension; and widespread gaps in access to health services for low- and middle-income people have not been resolved. Additionally, public health measures such as mask requirements that helped protect the unvaccinated have now been scaled back, raising the prospects that elevated mortality related to COVID-19 may continue. However, even if COVID-19 mortality were fully eliminated, the U.S. would still likely suffer over 600,000 excess deaths each year, with most occurring among Americans under 65 years. Preventing future Missing Americans will require policies that redress the consequences of structural racism(65) and bolster the economic and social determinants of population health.(66)

## Data Availability

All data produced are available online from the Human Mortality Database and CDC Wonder database.

https://www.mortality.org

https://www.wonder.cdc.gov

## Acknowledgments

None.

## Author contributions

Conceptualization: JB, DH, SW

Methodology: JB, AV, DH, SW

Investigation: JB

Visualization: JB, DH, SW

Interpretation: JB, AS, JR, AV, MB, DH, SW

Writing – original draft: JB

Writing – review & editing: JB, AS, JR, AV, MB, DH, SW

## Data and materials availability

All data used in the analysis are available from public sources. Code is available from the authors.

## Supplementary Materials

Materials and Methods

Tables S1 to S3

Figs. S1 to S5

## SUPPLEMENTARY MATERIALS

### Appendix A. Materials and Methods

#### Materials

##### Data sources

Data on deaths and population denominators were obtained from the Human Mortality Database (HMD),(27) which compiles official data from national vital registries and censuses. HMD is maintained by researchers at the Max Planck Institute for Demographic Research and the University of California-Berkeley.

The standard HMD mortality “long series” includes annual, age-specific mortality data (deaths, exposure time, and rates) for 41 countries, starting as early as 1751 (Sweden) and extending up to 2020 (for most countries) and to 2021 for Denmark and Sweden. These data are harmonized into standardized a format. During the COVID-19 pandemic, due to the demand for more real-time data on the mortality impact of the pandemic HMD has compiled and published provisional mortality data released by national vital statistics agencies. These data, published as the Short-Term Mortality Fluctuations (STMF) database include weekly death counts for 38 countries from the mid-2010s (for most countries) through early 2022. The STMF data are updated regularly. A limitation of the STMF database is that different countries report different age bands for provisional deaths. While HMD publishes harmonized age categories, these categories are very broad (e.g. 15-65 years is a single group). We utilized the original input data provided by countries and utilized in the STMF database.

The STMF data have limitations. Data may be incomplete due to lags between the date of death and registration, although such issues are mitigated as the STMF is updated. Data from the U.K. are reported based on date of registration rather than date of occurrence. Data from Canada exclude deaths in the Yukon (<0.1% of Canada’s population). Finally, Japan is not currently included in the STMF database, although its HMD “long series” runs through 2020.

##### Countries and country-years included in the analysis

We compared mortality trends in the U.S. with mortality trends in 18 other wealthy countries. The comparison set of countries included: Austria, Belgium, Canada, Denmark, Finland, France, Germany, Iceland, Italy, Japan, Luxembourg, Netherlands, Norway, Portugal, Spain, Sweden, Switzerland, and United Kingdom. These countries represented <u>all countries with mortality data available from HMD beginning in 1960 or earlier and ending in 2020 or later</u>, after excluding former-Soviet/Eastern Bloc nations and two countries (Australia, New Zealand) with incomplete data.

Details on the panel of countries are provided in **Table S1**. Our analysis includes all country-years from 1933, the first year of U.S. data, through 2021. All countries were observed starting in 1933 with the exception Portugal (1940), Austria (1947), Japan (1947), Germany (1956), and Luxembourg (1960). The Germany series combines East and West Germany prior to 1989. All countries were observed through 2021, with the exception of Japan, for which we carried forward 2020 ASMRs to 2021. Data for Canada were incomplete in 2021 and missing weeks were imputed by assuming that the average mortality rate in prior weeks prevailed for the rest of the year.

##### Data extraction and manipulation

For each country, we extracted annual age-specific mortality rates, deaths, and denominators (exposure time) for 5-year age groups from the HMD long series. We then extracted data on deaths for the most recent years using the original input data from the STMF. Data were downloaded on June 7, 2022. We aggregated the STMF weekly data to annual counts. Because different countries reported deaths for different age bands in the STMF, we allocated deaths within STMF age groups to 5-year age groups based on the distribution of deaths in the last year available from the HMD long series. For example, in the STMF, Germany reports deaths for 5-year age groups among persons 30 and over but collapses all ages under 30 years. Using the distribution of deaths in 2017 – Germany’s last year in the “long series” – we reallocated deaths under 30 years to ages 0-4, 5-9, …, 25-29 for 2018-2021. Denominators to match the STMF death counts were obtained by fitting a linear trend to the last five years of exposure data available from the HMD long series for each 5-year age group in each country (e.g. Germany, ages 40-44, 2013-2017) to obtain predictions for the most recent years (e.g. Germany, ages 40-44, 2018-2021).

Our final dataset consisted of annual age-specific mortality rates by 5-year age group, for 19 countries (U.S. and 18 “other wealthy nations”) with data from 1933 through 2021. We assessed the accuracy of the STMF data by comparing the STMF and HMD estimates for country-years with overlapping data between the two datasets. ASMRs were closely similar **(Table S1)**. Finally, to avoid false precision in reporting, we aggregated deaths and exposure time to the ten-year age bands reported by the U.S. in the STMF data: 0-4, 5-14, 15-24, …, 85+ years. Additionally, for our age-stratified analyses, we constructed six wider age groups to facilitate visual and tabular presentation of the data: 0-14, 15-44, 45-64, 65-74, 75-84, 85+.

As a benchmark against which to compute “excess” U.S. mortality, we calculated the average ASMR across “other wealthy nations” for each age group and year (excluding countries whose data were not available for that year). We weighted countries by population (following (11)), and compared the mortality rates of U.S. residents to the weighted average mortality rates of residents of other wealthy nations.

We also calculated age-specific mortality for U.S. racial and ethnic population groups for 1999-2021 based on mortality figures and midyear population denominators stratified by age and race/ethnicity extracted from the Multiple Cause Mortality Files hosted on the CDC WONDER database.(28) (1999 was the first year of the MCMF database on CDC WONDER.) Data were downloaded April 14, 2022. We computed ASMRs for: Non-Hispanic Black (henceforth Black), Non-Hispanic White (White), Hispanic, Non-Hispanic Asian/Pacific Islander (Asian/Pacific Islander), and Non-Hispanic American Indian/Alaskan Native (Native American). For each population subgroup, ASMRs were computed for ten-year age groups, aligned with the data from HMD. For ease of exposition, we report on the wider age bands described above.

#### Methods

For each year, 1933-2021, we compared U.S. ASMRs for each age group (0-14, 15-44, 45-64, 65-74, 75-84, 85+) with the ASMRs of people residing in 18 “other wealthy nations” (OWN). To assess excess mortality on a relative scale, we computed ASMR ratios for each year, 1933-2020, comparing the U.S. with the OWN benchmark. We additionally computed ratios comparing ASMRs for U.S. racial and ethnic population groups with the ASMRs of OWN, each year 1999-2021.

To assess excess mortality on an absolute scale, we calculated the number of deaths that would have been observed in the U.S. in each year if the U.S. had the ASMRs of the OWN (multiplying the U.S. population distribution by OWN ASMRs). We then subtracted this number from the number of observed deaths in the U.S. to compute “excess deaths”.

In addition to excess deaths, we computed “years of life lost” (YLL) which weights deaths by the number of years that a person would have been expected to live had they survived. Following (11), we compute YLL weighting deaths by the U.S. age-specific life expectancy in that year.

Finally, to compare changes in mortality associated with the COVID-19 pandemic, we computed changes in age-specific mortality rates from 2019 to 2020 and from 2019 to 2021 for the total U.S. population, for U.S. racial and ethnic groups, and for the comparison group of OWN.

### Appendix B. Supplementary Figures

**Fig. S1.**
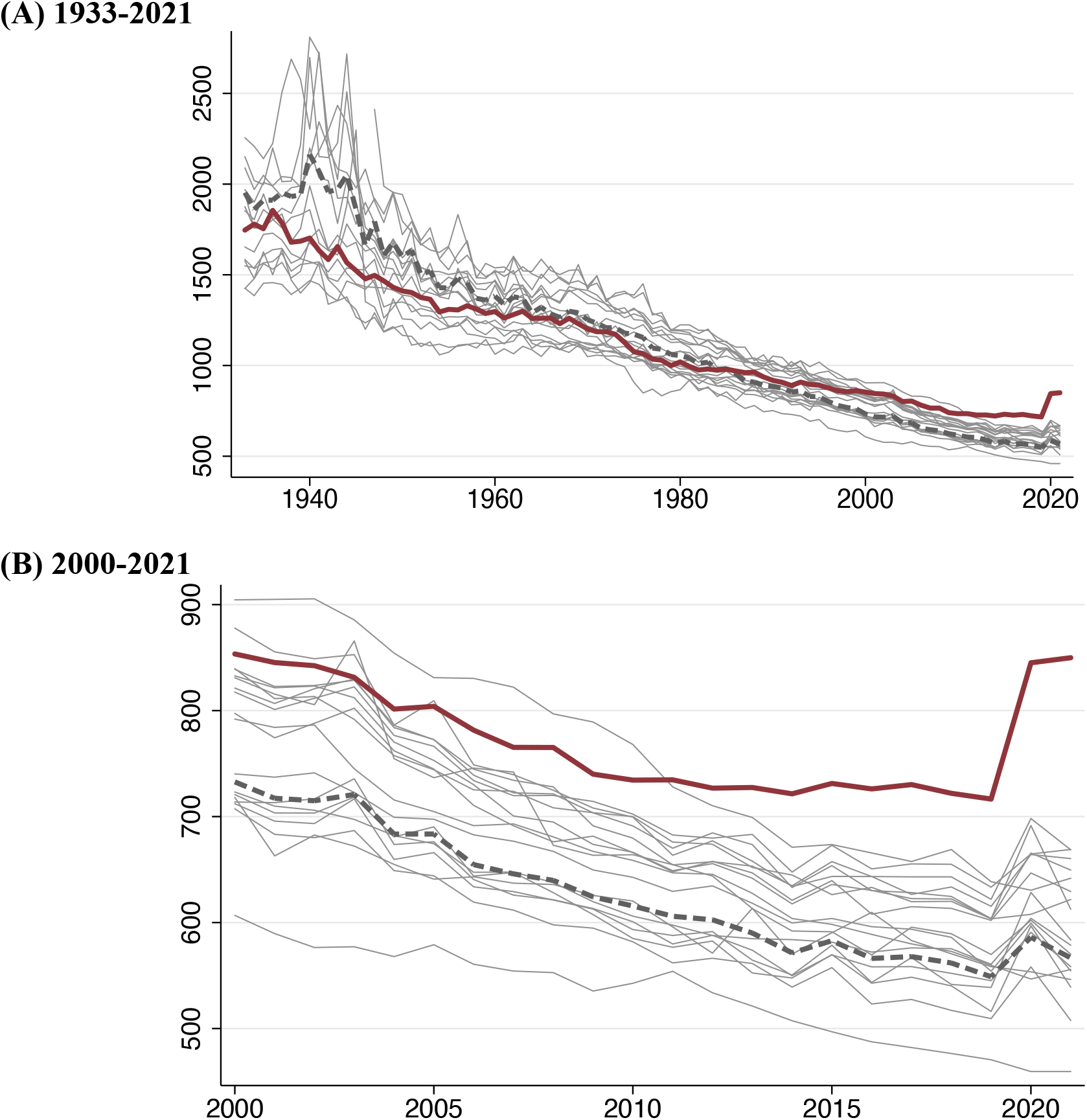
Age-standardized mortality trends in the U.S. and other wealthy nations. Figure shows deaths per 100K population, (A) 1933-2021 and (B) 2000-2021. Dark red line is U.S.; pink line is the population-weighted average of other wealthy nations; grey lines are country-specific trends for non-U.S. nations. Total mortality was age-standardized to the 2000 U.S. population age distribution.

**Fig. S2.**
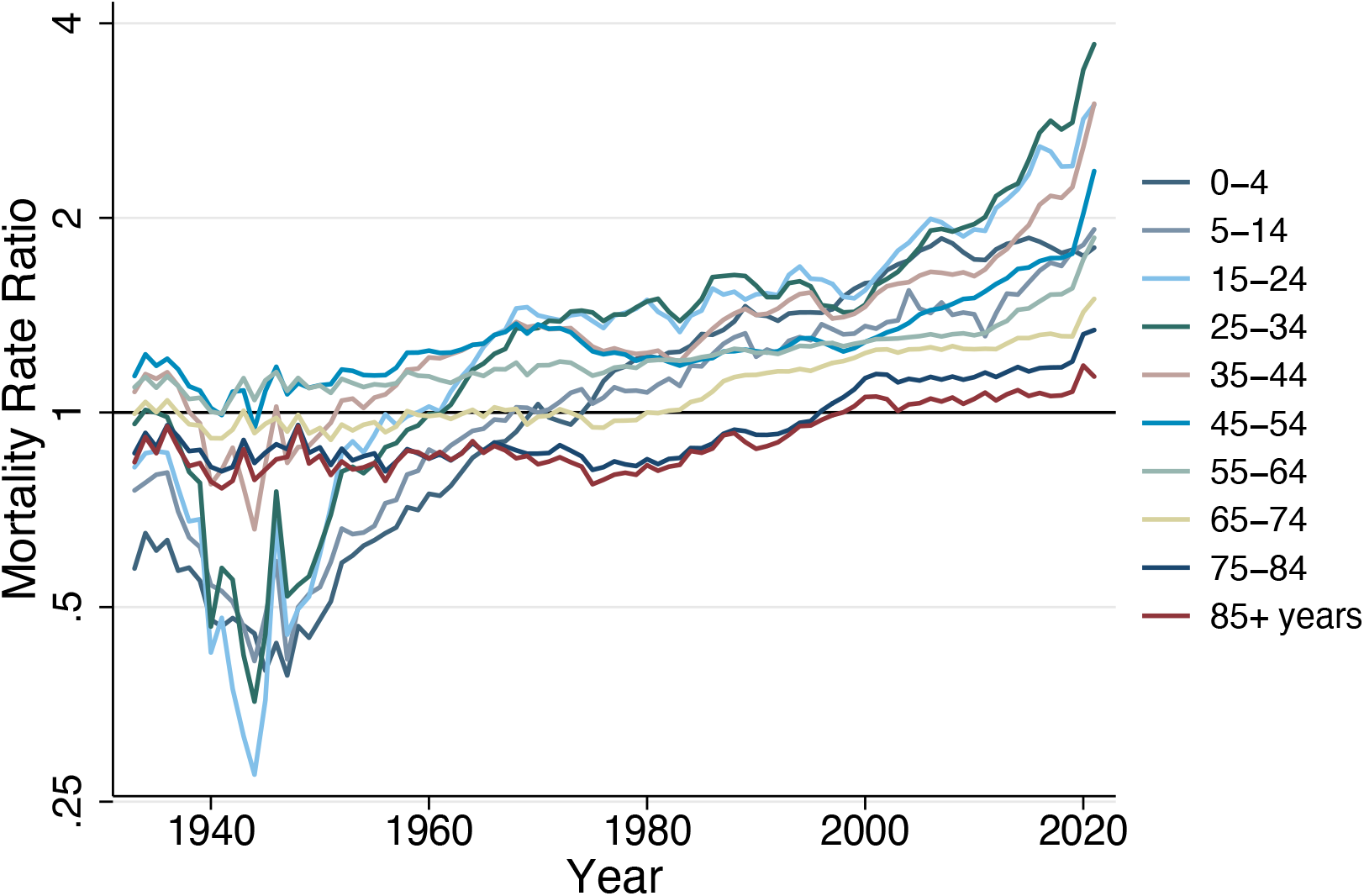
Ratio of U.S. mortality to mortality in other wealthy nations, by age group. Note: Figure shows mortality rate ratios for each age group, 1933-2021. The average of other wealthy nations excludes Portugal prior to 1940, Austria and Japan prior to 1947, Germany prior to 1956, and Luxembourg prior to 1960. From 1960, all countries are represented.

**Fig. S3.**
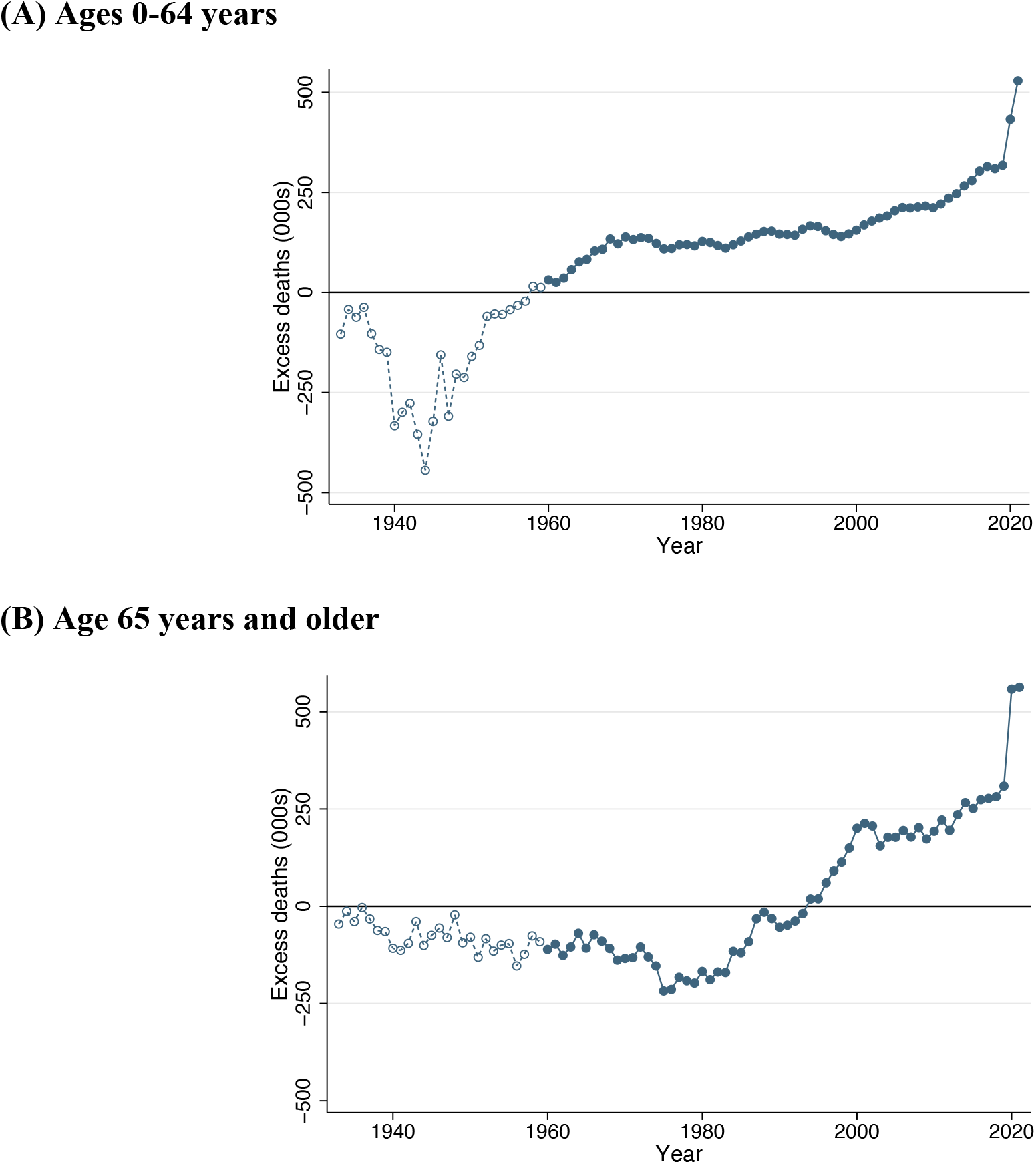
Missing Americans, 1933-2021, under 65 years vs. 65 and older. Note: For each age range, figure shows difference between observed deaths in the U.S. and the number of deaths that would have occurred in the U.S. if the U.S. had age-specific mortality rates equal to the average of other wealthy nations. The average of other wealthy nations excludes Portugal prior to 1940, Austria and Japan prior to 1947, Germany prior to 1956, and Luxembourg prior to 1960. From 1960, all countries are represented.

**Fig. S4.**
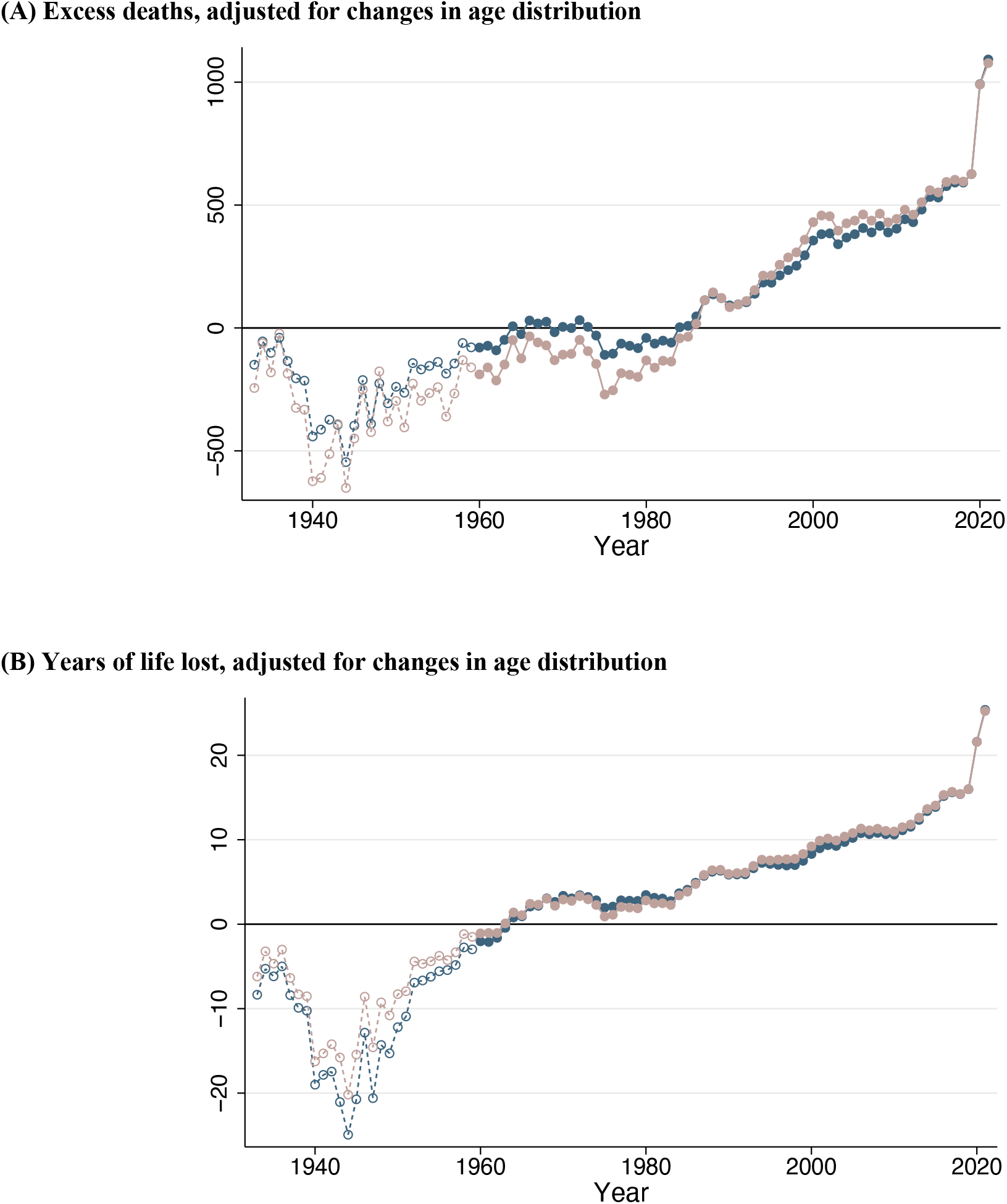
Excess deaths and years of life lost, 1933-2021, standardized by U.S. 2019 age distribution. Note: Figure shows the difference between observed deaths in the U.S. and the number of deaths that would have occurred in the U.S. if the U.S. had age-specific mortality rates equal to the average of other wealthy nations. Mortality rates are standardized to the 2019 U.S. age distribution, while preserving the total U.S. population in each year. The average of other wealthy nations excludes Portugal prior to 1940, Austria and Japan prior to 1947, Germany prior to 1956, and Luxembourg prior to 1960. From 1960, all countries are represented.

**Fig. S5.**
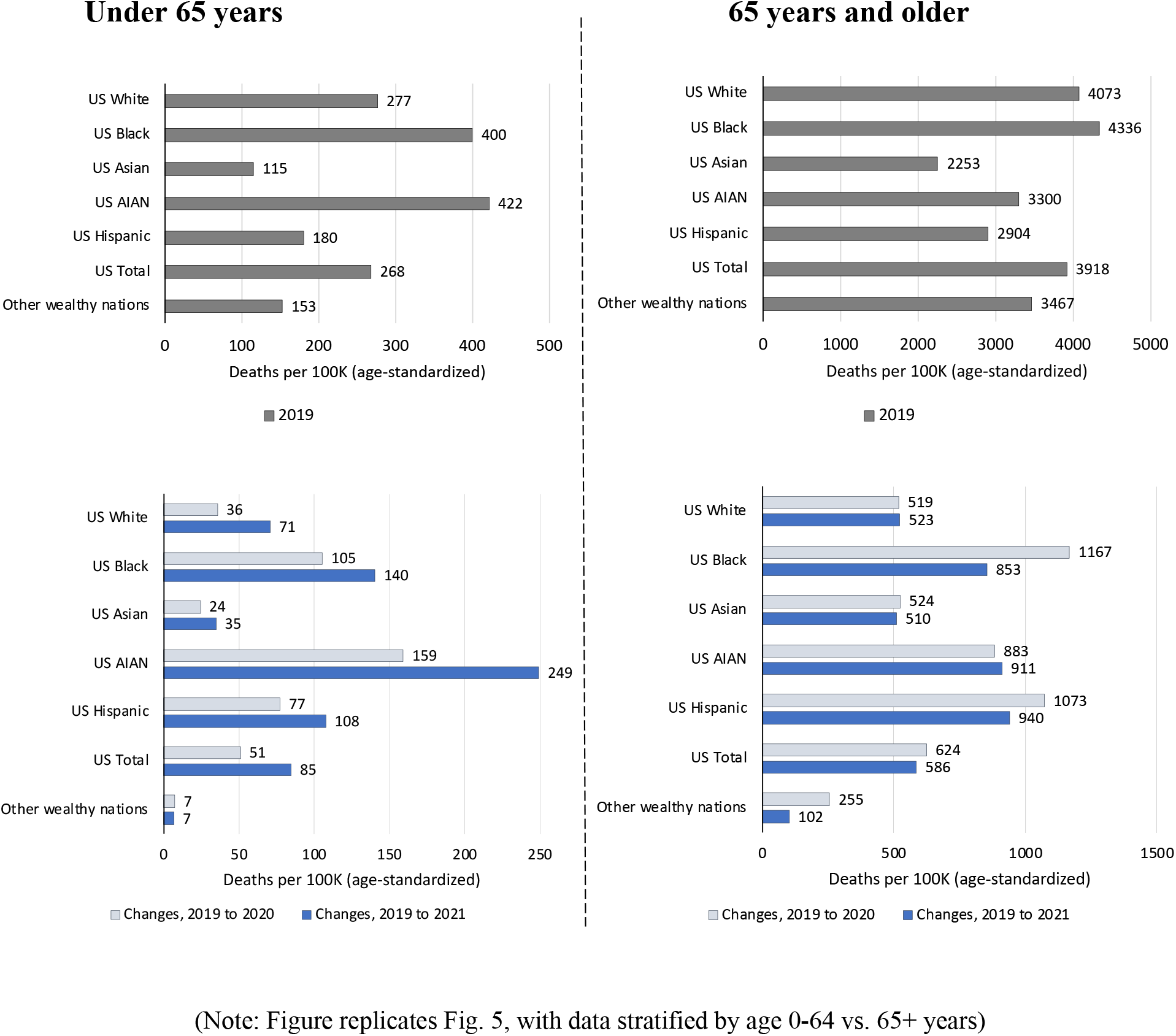
Under-65 and 65 years and older age-standardized mortality rates for U.S. racial and ethnic groups and the average of other wealthy nations: 2019-2021. Source: Multiple Cause Mortality Files from CDC Wonder and Human Mortality Database. Note: Figure shows data stratified by age: under 65 years (left) and 65 years and older (right). Top panels show 2019 mortality rates and bottom panels show changes in mortality 2019 to 2020 and 2019 to 2021. Mortality rates are standardized to the 2019 U.S. population age distribution. Figure compares mortality rates for U.S. racial/ethnic groups: Hispanic, Non-Hispanic White (White), Non-Hispanic Black (Black), Non-Hispanic Asian/Pacific Islander (Asian), Non-Hispanic American Indian/Alaskan Native (AIAN).

### Appendix C. Supplementary Tables

**Table S1.** Panel of countries included in the analysis.

| Country | Country Code | Start year (HMD long) | End year (HMD long) | End year (STMF) | % Diff: HMD long vs. STMF | Population (2020) |
| --- | --- | --- | --- | --- | --- | --- |
| Austria | AUT | 1947 | 2019 | 2021 | -2.3% | 9,010,743 |
| Belgium | BEL | 1933 | 2020 | 2021 | 0.1% | 11,512,420 |
| Canada | CAN | 1933 | 2019 | 2021 | -0.2% | 38,394,153 |
| Switzerland | CHE | 1933 | 2020 | 2021 | 0.1% | 8,707,675 |
| Denmark | DNK | 1933 | 2021 | N/A | N/A | 5,856,866 |
| Spain | ESP | 1933 | 2020 | 2021 | 0.1% | 47,568,022 |
| Finland | FIN | 1933 | 2020 | 2021 | 0.2% | 5,538,503 |
| France | FRA | 1933 | 2019 | 2021 | -0.3% | 65,397,615 |
| Germany | GER | 1956 | 2017 | 2021 | 0.1% | 84,894,520 |
| Iceland | ISL | 1933 | 2020 | 2021 | 0.1% | 375,469 |
| Italy | ITA | 1933 | 2019 | 2021 | 1.2% | 59,487,834 |
| Japan | JPN | 1947 | 2020 | N/A | N/A | 123,423,636 |
| Luxembourg | LUX | 1960 | 2020 | 2021 | 0.2% | 642,874 |
| Netherlands | NLD | 1933 | 2019 | 2021 | -0.3% | 17,540,875 |
| Norway | NOR | 1933 | 2020 | 2021 | 0.0% | 5,418,375 |
| Portugal | PRT | 1940 | 2020 | 2021 | 0.3% | 10,277,536 |
| Sweden | SWE | 1933 | 2021 | N/A | N/A | 10,417,555 |
| United Kingdom | UK | 1933 | 2020 | 2021 | 0.0% | 67,173,059 |
| United States | USA | 1933 | 2020 | 2021 | 0.4% | 332,632,365 |

Note: Demographic data were obtained from the Human Mortality Database (HMD). We extracted data both from the HMD long series (HMD long) and from the HMD short term mortality fluctuations dataset (STMF). We included all countries represented in the HMD that had data starting in 1960 or earlier and extending to 2020 or later. We excluded former Soviet and Eastern Bloc countries (with the exception of East Germany, which was combined with West Germany pre-1989), and we excluded Australia and New Zealand, which had incomplete STMF data. Percent difference (% diff) between HMD long and STMF reports the percent difference in total number of deaths in the two databases for those country-years where both were represented. Denmark and Sweden had long-series data through 2021 and STMF data were not used for those countries. Japan did not have STMF data.

**Table S2.** Excess mortality in the U.S. relative to other wealthy nations: 2019-2021.

| Year | Age | Deaths per 100K |  | Total Deaths |  | Excess deaths |  |  |
| --- | --- | --- | --- | --- | --- | --- | --- | --- |
|  |  | USA | Other wealthy | USA observed | Counterfactual | Ratio | Difference | % Excess |
| 2019 | 0-64 years | 268 | 153 | 737,393 | 419,541 | 1.76 | 317,852 | 43% |
|  | 65+ years | 3,962 | 3,384 | 2,117,426 | 1,808,926 | 1.17 | 308,500 | 15% |
|  | *Total* | 869 | 678 | 2,854,820 | 2,228,467 | 1.28 | 626,353 | 22% |
| 2020 | 0-64 years | 317 | 160 | 874,247 | 440,859 | 1.98 | 433,388 | 50% |
|  | 65+ years | 4,595 | 3,572 | 2,509,477 | 1,950,997 | 1.29 | 558,480 | 22% |
|  | *Total* | 1,023 | 723 | 3,383,724 | 2,391,856 | 1.41 | 991,868 | 29% |
| 2021 | 0-64 years | 351 | 160 | 969,709 | 440,863 | 2.20 | 528,846 | 55% |
|  | 65+ years | 4,419 | 3,418 | 2,485,895 | 1,922,449 | 1.29 | 563,447 | 23% |
|  | *Total* | 1,039 | 710 | 3,455,604 | 2,363,311 | 1.46 | 1,092,293 | 32% |
| Change,<br>2019-2020 | 0-64 years | 48 | 7 | 136,854 | 21,318 | 6.42 | 115,536 | 84% |
|  | 65+ years | 633 | 188 | 392,051 | 142,071 | 2.76 | 249,980 | 64% |
|  | *Total* | 154 | 45 | 528,905 | 163,389 | 3.24 | 365,516 | 69% |
| Change,<br>2019-2021 | 0-64 years | 83 | 7 | 232,316 | 21,322 | 10.90 | 210,994 | 91% |
|  | 65+ years | 458 | 33 | 368,469 | 113,522 | 3.25 | 254,946 | 69% |
|  | *Total* | 170 | 32 | 600,784 | 134,844 | 4.46 | 465,940 | 78% |

**Table S3.** U.S. excess deaths relative to alternative international comparators: 2019-2021.

| Year | Age | **U.S. Excess Deaths Relative to International Comparator Consisting of:** |  |  |
| --- | --- | --- | --- | --- |
|  |  | 18 Other Wealthy Nations<br>(Main Specification) | 6 Other "Group-of-7" Nations<br>(Woolhandler, et al 2021) | 5 European Nations<br>(Preston-Vierboom 2021) |
| 2019 | 0-64 years | 317,852 | 311,560 | 295,154 |
|  | 65+ years | 308,500 | 327,200 | 213,113 |
|  | Total | 626,353 | 638,760 | 508,267 |
| 2020 | 0-64 years | 433,388 | 425,799 | 403,623 |
|  | 65+ years | 558,480 | 599,025 | 371,948 |
|  | Total | 991,868 | 1,024,823 | 775,570 |
| 2021 | 0-64 years | 528,846 | 519,751 | 498,860 |
|  | 65+ years | 563,446 | 586,649 | 412,663 |
|  | Total | 1,092,293 | 1,106,400 | 911,523 |
| Change,<br>2019-2020 | 0-64 years | 115,536 | 114,239 | 108,469 |
|  | 65+ years | 249,980 | 271,825 | 158,835 |
|  | Total | 365,516 | 386,064 | 267,304 |
| Change,<br>2019-2021 | 0-64 years | 210,994 | 208,191 | 203,706 |
|  | 65+ years | 254,946 | 259,449 | 199,550 |
|  | Total | 465,940 | 467,640 | 403,256 |

